# Activation of the Venous Muscle Pump by Neuromuscular Stimulation of the Common Peroneal Nerve Reduces Postoperative Edema in the Foot and Ankle

**DOI:** 10.1101/2025.05.20.25327913

**Authors:** Andrea Sallent, Nicholas Abidi, Albert Baduell, Shelain Patel

## Abstract

Edema after foot and ankle surgery is associated with unsatisfactory outcomes. It impairs postoperative mobilization, range of motion, pain, wound healing, and risk of venous thrombosis. The aim of this study is to determine if activation of the venous muscle pump by neuromuscular stimulation of the common peroneal nerve reduces postoperative edema in the foot and ankle.

80 patients having forefoot or hindfoot surgery were randomized to receive standard care following surgery, or a wearable self-adhesive neuromuscular stimulation device applied to the skin below the knee laterally at the head of fibula. This provided intermittent (1 hertz) stimulation of the common peroneal nerve, eliciting a twitch of the muscles of the leg. Edema was measured as the difference between the involved and uninvolved limb, measured by figure-of-eight method of the foot and ankle. Baseline was measured preoperatively and then 14 days following surgery.

Figure-of-eight measurement in patients receiving standard care increased relative to baseline by 4.9% (95% CI 4.3%-5.4%) 14 days post-surgery, whereas patients receiving neuromuscular stimulation increased by 3.3% (95%CI 2.7%-3.6%). Patients receiving neuromuscular stimulation had 33% less edema (p<0.05) than those receiving standard care.

## Introduction

Complications following foot and ankle surgery are common (1). Among these, one of the most frequently reported is edema (2), which has sequalae of other poor outcomes after ankle surgery (3), including impaired wound healing (4).

Postoperative edema has a negative impact on pain and functional recovery, reducing mobility, well-being and quality of life; furthermore, it impedes rehabilitative progress, and extends length of stay (5-7).

Not treated promptly, chronic progression of edema can lead to disablement, based on secondary posttraumatic lymphedema (8-9). Almost half of ankle surgery patients report edema at 5 years (10).

Interventions for postoperative edema include compression stockings applied locally. These have been shown to reduce edema and so improve healing outcomes but are also associated with a higher rate of infection (11). Another treatment offered is intermittent pneumatic compression (IPC), which appears to be effective, but is cumbersome and suffers from poor patient concordance (12). The benefits of compression are also weighed against the risks of compartment syndrome, nerve palsy, blistering, and necrosis (13-14).

A novel treatment comprises intermittent neuromuscular electrostimulation (NMES) of the common peroneal nerve at 1 hertz (Hz), to elicit a muscular twitch, so activating the venous muscle pumps of the leg and foot. The geko^®^ device (Firstkind Ltd, Daresbury, Cheshire, United Kingdom) has been previously shown to increase venous output from the leg (15-17) and has been shown to reduce edema preoperatively in ankle fracture (18). It has the advantage over conventional or intermittent compression that it is applied remotely to the surgical site (Fig. 1).

**Figure 1.**
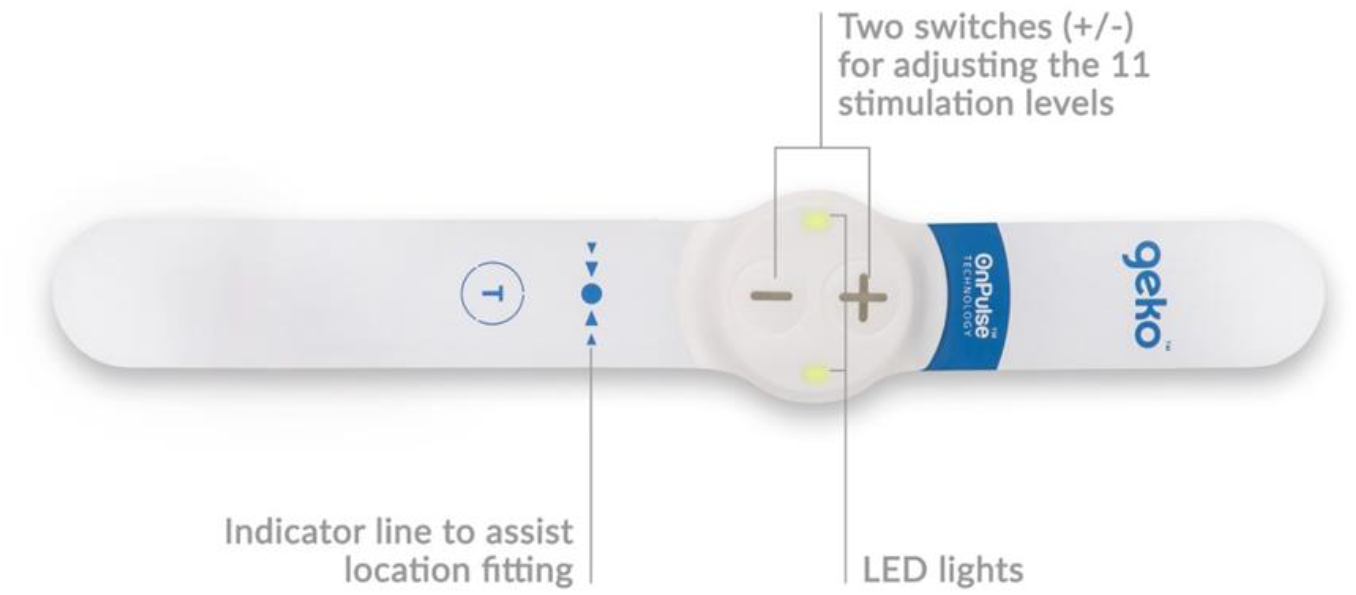
The geko® Device, Firstkind Ltd, Daresbury, Cheshire, United Kingdom. The device includes an indicator line to assist with accurate anatomical placement, two control switches (±) for adjusting among 11 stimulation levels, and LED lights that indicate operational status. The device delivers NMES via OnPulse® technology to activate the common peroneal nerve.

## Patients and Methods

### Aims

The primary aim of this study was to evaluate whether activation of the venous muscle pump via NMES of the common peroneal nerve reduced postoperative edema in the foot and ankle. Secondary outcome measures included the incidence of adverse events, pain intensity assessed using the Visual Analogue Scale (VAS), and surgical site healing.

Ethical approval was obtained from the appropriate ethics committees in Spain, the United States, and the United Kingdom, as the study was conducted at sites within these countries. The trial was registered on ClinicalTrials.gov (Identifier: NCT04927234) and conducted in accordance with national ethical guidelines, the Declaration of Helsinki, EN ISO 14155:2020, and ICH E6 (R2) Good Clinical Practice. All relevant legal and regulatory requirements were met. Study design and reporting adhered to CONSORT guidelines (19).

### Methods

This study was an open label, multi-center, prospective randomized study. A schematic of the study design and patient pathway is presented in Figure 2. Participants were identified to undergo forefoot surgery: hallux valgus correction; 1st metatarsophalangeal joint (MTPJ) cheilectomy; MTPJ cheilectomy; 1st MTPJ fusion; lesser toe surgery; Morton’s neuroma excision; and/or hindfoot surgery: ankle fusion; ankle replacement; ankle arthroscopy; mid/hindfoot fusion.

**Figure 2.**
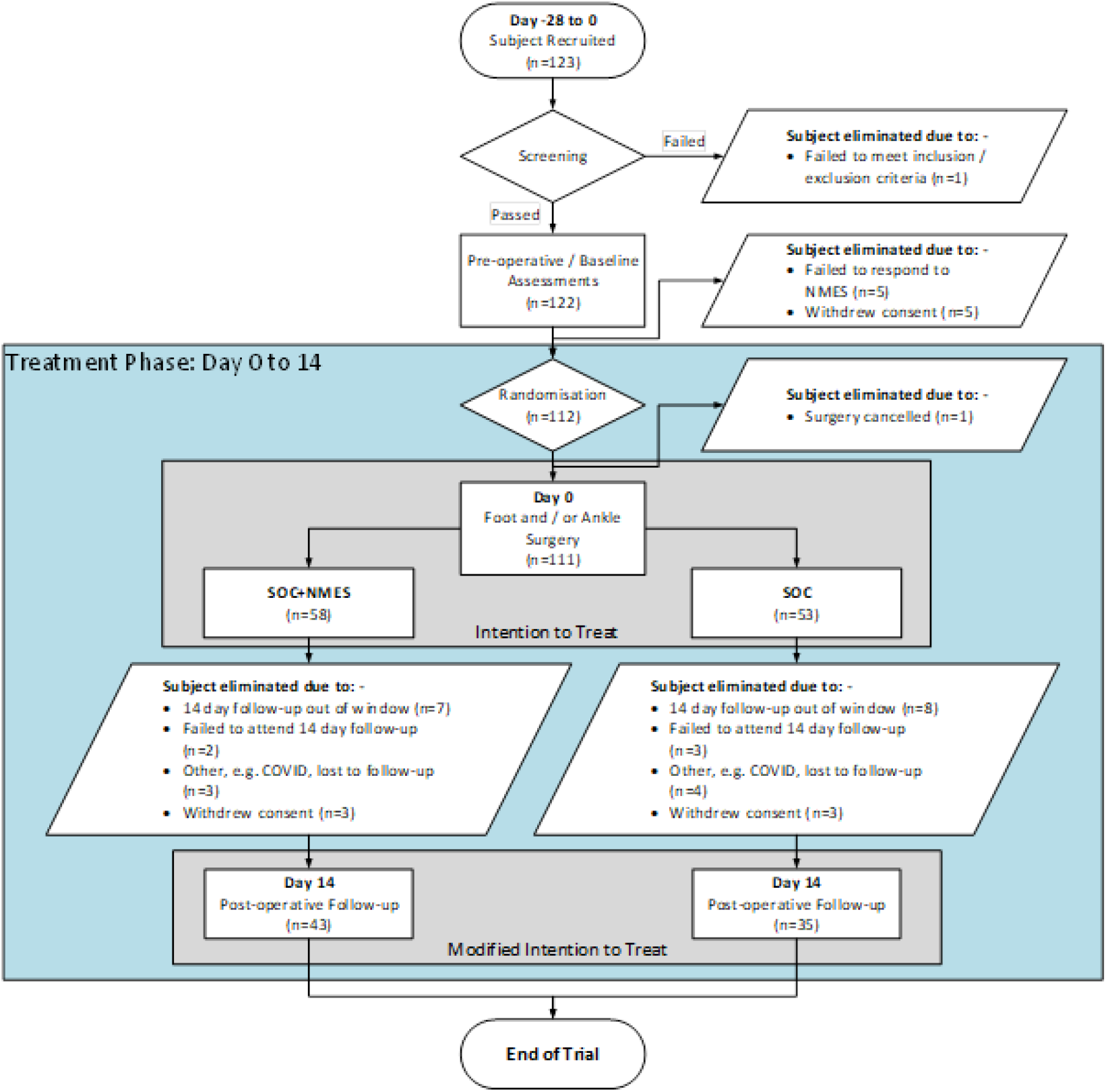
Flow Diagram of Participant Progression through the Study. A total of 123 participants were recruited and screened, with 112 undergoing randomization and 111 foot and/or ankle surgery (Day 0). Participants were assigned to either the intervention group (SoC + NMES, N=58) or standard of care (SoC, N=53). Reasons for elimination from the modified intention-to-treat population are detailed for each group, including follow-up outside the protocol window, failure to attend, withdrawal of consent, or loss to follow-up. Post-operative follow-up at Day 14 was completed by 43 participants in the SoC + NMES group and 35 participants in the SoC group.

### Inclusion Criteria

- Aged ≥ 18 years
- No more than 2 falls in the preceding 12 months
- Intact healthy skin at the site of NMES device application
- Listed for forefoot and / or hindfoot surgery
- Understand and willing to participate in the study and can comply with study procedures
- Willing and able to give written informed consent

### Exclusion Criteria

- Pregnant
- Use of any other neuro-modulation device
- Trauma to the lower limbs that would prevent NMES from stimulating the common peroneal nerve

### Sample Size Determination

Based on the mean and standard deviation difference between study arms in previous studies (20) it was determined that 50 patients would be needed in each group for a power of 95% and type I error (alpha) of 0.05. Calculation based on the formula:

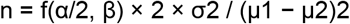

where μ1 and μ2 are the mean outcome in the control and experimental group respectively, σ is the standard deviation. Assuming an attrition rate of 18% of patients, 61 patients were scheduled to be recruited into each arm of the trial.

Participants were randomized on a 1:1 basis using the Castor Electronic Data Capture (EDC) system, which employed a computer-generated randomization schedule to allocate participants to the intervention or control group. The intervention group received NMES therapy in addition to standard care (SoC) and the control group received SoC only. SoC consisted of pain relief (non-steroidal anti-inflammatory drugs), elevation, and off-loading.

### Endpoints

Edema was quantified using the Figure-of-Eight Method (FO8) for measuring ankle joint swelling (21). FO8 has been shown to correlate well to water displacement methods (22-23) and to optical techniques (24), demonstrating reliability (intraclass correlation coefficient 0.94) and validity (r = 0.65; *p* < 0.001) (25), with considerable advantages in terms of convenience in a clinical setting (26-27). The FO8 method is illustrated in Figure 3.

**Figure 3.**
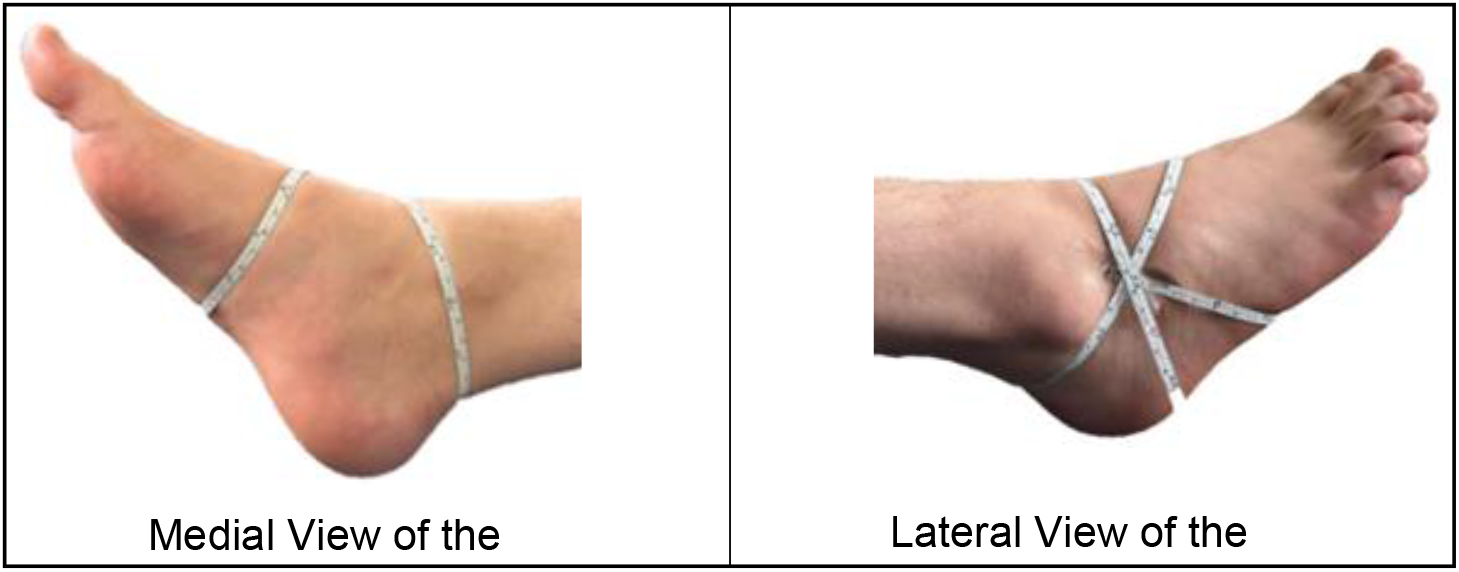
Medial and Lateral Views of the Foot Demonstrating the Figure-of-Eight Measurement Method. Key bony landmarks were marked with an indelible pen to ensure reproducibility, and a disposable paper tape was used to trace the figure-of-eight pattern.

Participants were supine with both feet extended beyond the end of the examination table to the level of midcalf with the ankle in neutral position of eversion/inversion in 90° of dorsiflexion. The following bony landmarks were marked with an indelible pen to allow reproducibility and then connected by doing a figure-of-eight using a disposable paper tape.

- Beginning midway between tibialis anterior tendon and lateral malleolus
- Medially across instep just distal to navicular tuberosity.
- Across arch to base of 5th metatarsal, then across tibialis anterior tendon
- Around ankle joint, just distal to tip of medial malleolus
- Across Achilles tendon, just distal to lateral malleolus, back to starting point.

Each measurement was repeated 3 times on the involved limb and the un-involved limb, and the mean calculated.

Secondary endpoints included the incidence of adverse events, pain intensity assessed using VAS, and the status of surgical site healing as evaluated at follow-up.

### Intervention

The NMES device (geko^®^ T device, Firstkind Ltd, Daresbury, Cheshire, United Kingdom) has 11 stimulation settings designed to ensure activation of the venous muscle pumps of the calf and foot irrespective of individual skin impedance. The optimal setting varies between patients and is achieved when the setting selected is enough to activate an intermittent (1 Hz) contraction of the leg muscles, observable as a visible dorsiflexion/eversion twitch of the foot, while remaining comfortable to the patient.

The NMES device was fitted only to the operated leg as per the fitting instructions just below the knee, to the skin over the fibular head. NMES was delivered for the first 24 hours post-surgery, followed by 12 hours/day at home until the first post-operative follow-up visit (FU1), at Day 14 ± 2 days post-surgery.

## Results

### Demographics

Table 1 shows the demographic breakdown of the intervention group compared with the SoC group, presenting mean and standard error (SE) for each parameter

**Table 1.** Baseline Demographic Characteristics of Participants in the Intervention and Standard of Care (SoC) Groups. Values for age and BMI are presented as means ± standard error (SE), with p-values from independent t-tests. Gender is reported as number of participants per group; no statistical comparison was performed for categorical variables (N=111).

|  | Intervention | SoC | Intervention | SoC | t-test |
| --- | --- | --- | --- | --- | --- |
|  | Average | Average | SE | SE |  |
| **Age** | 52.03 | 53.79 | 1.89 | 2.18 | 0.54 |
| **BMI** | 27.37 | 29.23 | 0.66 | 0.93 | 0.11 |
| **Males (n)** | 14 | 16 |  |  |  |
| **Females (n)** | 44 | 37 |  |  |  |

The groups were similar in terms of age and body mass index (BMI). No statistically significant differences were found between the two groups by Student’s t-test. The two groups also had similar proportions of males to females, with no statistically significant differences between groups by chi-square test.

Figure 4 shows a breakdown of the participants by surgical procedure in the intervention group (A) and the standard care group (B). The participants underwent a miscellany of procedures including various fracture, scarf akin procedures, reductions/osteosyntheses flatfoot correction, Weil’s osteotomy, and forefoot reconstructions. No significant differences (Fisher’s exact test) were found between the intervention group and SoC group in terms of procedure frequencies.

**Figure 4.**
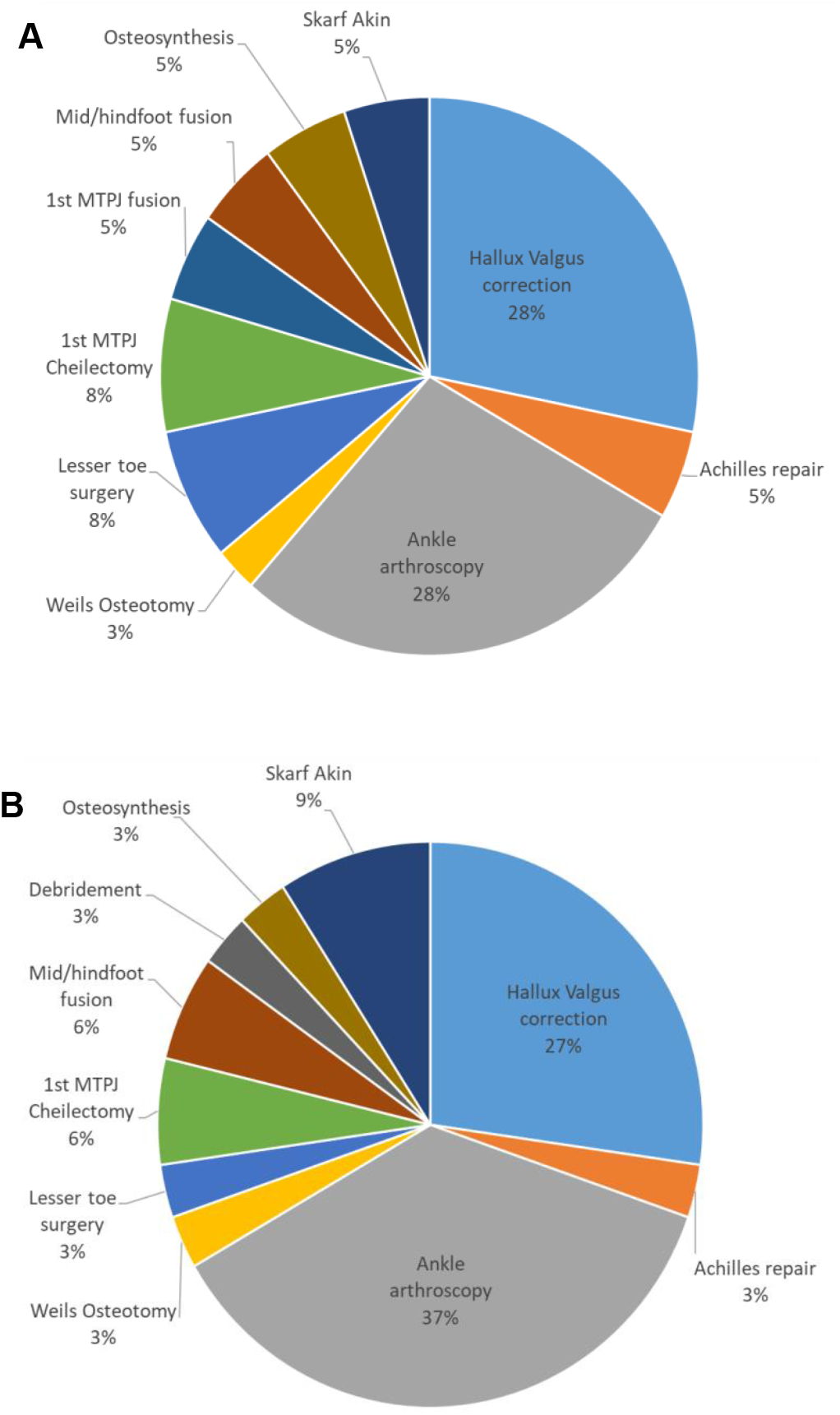
Breakdown of Surgical Procedures Performed in each Study Group. (A) Breakdown of surgeries performed in the intervention group (N = 58), (B) Breakdown of surgeries performed in the standard care group (N = 53). Percentages reflect the proportion of each procedure type within the respective group.

### Edema

Figure 5 shows the percentage difference in FO8 measurement between the involved and uninvolved legs at baseline, and at follow-up (FU1). In both the intervention group (blue) and the control group (orange), involved and uninvolved legs show similar measurements preoperatively (baseline). At FU1, both groups show a significant swelling of the involved leg compared to the uninvolved leg, with the intervention group showing a discrepancy of 3.2% (95% CI 2.7%-3.6%) and the control group showing a discrepancy of 4.9% (95% CI 4.3%-5.4%). This demonstrates a significant (p=0.045, un-paired t-test) protective effect in the intervention group.

**Figure 5.**
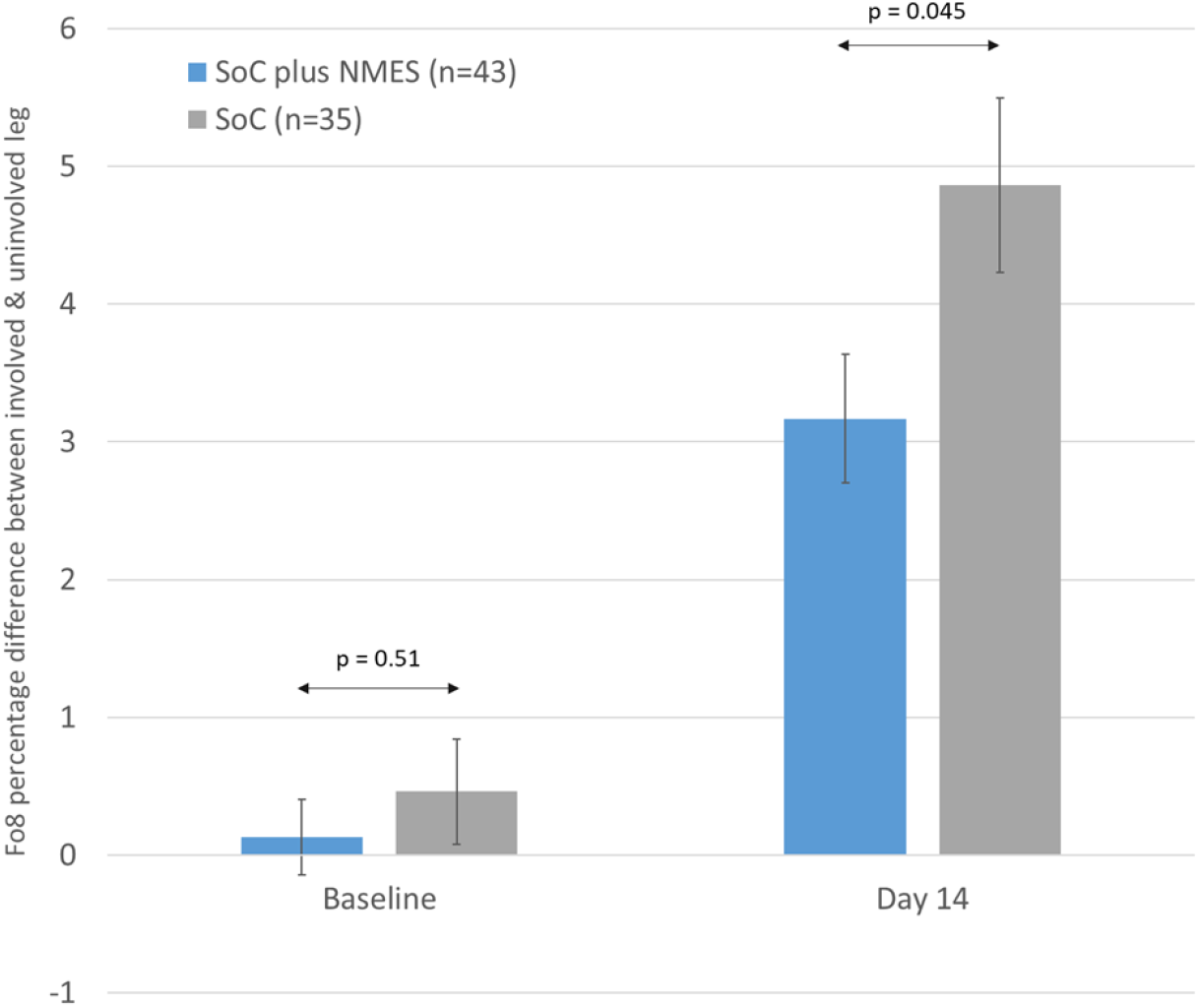
Figure-of-Eight Percentage Difference between the Involved and Uninvolved Legs at Baseline and Follow-up 1 (FU1) for the Intervention Group (N=43) and the Standard Care (SoC) Group (N=35). At baseline, there was no significant difference between groups (p = 0.51). By FU1, a statistically significant difference was observed (p = 0.045), with the SoC group exhibiting a greater percentage difference (4.86 ± 0.63%) compared to the intervention group (3.17 ± 0.47%)

### Secondary Endpoints

Although this study was not powered to perform inferential statistics on any of the secondary endpoints, results are presented descriptively below.

### Adverse Events

Figure 6 shows the incidence of adverse events in each group, presented as number of events per 100 patient days. The adverse events are categorized into non-device-related, non-serious adverse events (AE), device related non-serious adverse events (ADE), non-device-related serious adverse events (SAE), and device-related serious adverse events (SADE). There were no SAEs or SADEs in either group. The incidence of AE events was broadly similar in the two groups, at just over 1 event per 100 patient days. The intervention group saw a similar modest number of ADEs, which were related to skin redness at the device application site. It is worth noting that the manufacturer has recently introduced a new hydrogel adhesive on the device, which appears to ameliorate this skin redness issue. The absence of any ADEs in the SoC group is unremarkable, as no devices were used, and this designation is not applicable.

**Figure 6.**
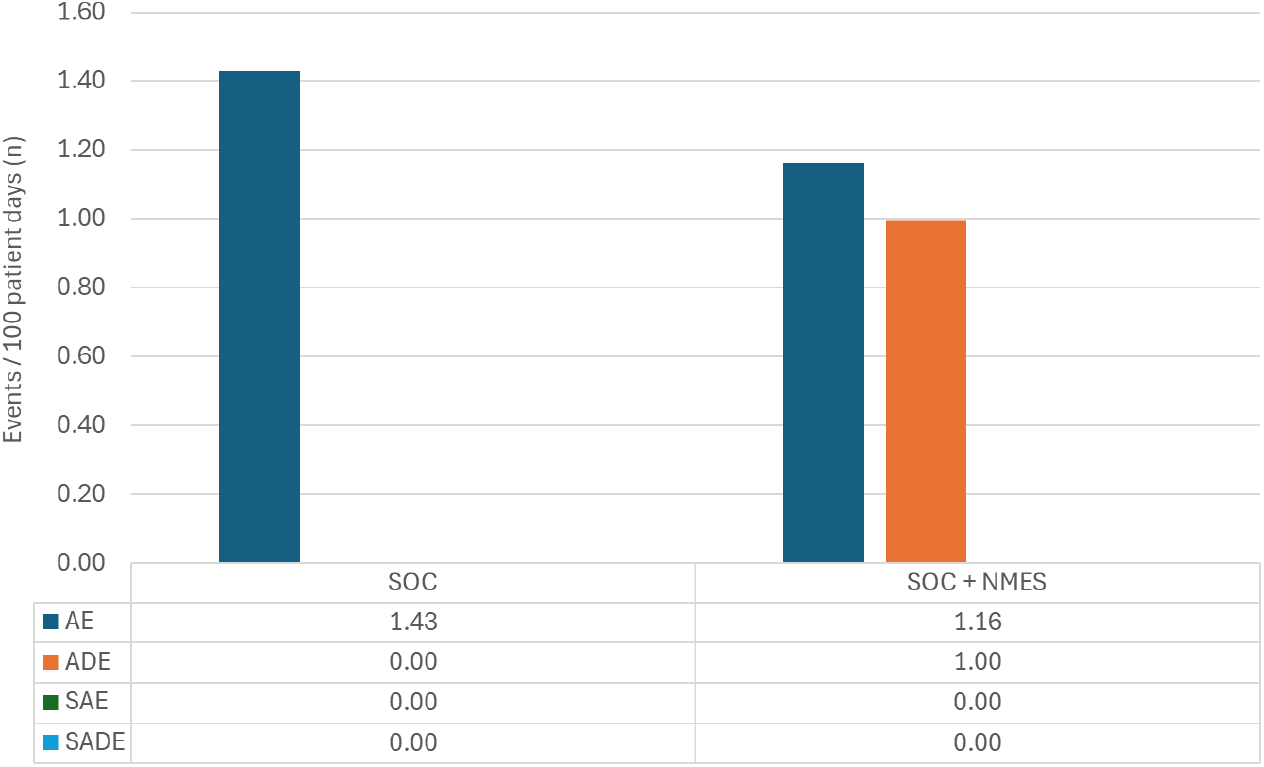
Incidence of Adverse Events per 100 Patient Days across the Two Treatment Groups (N=78). The rate of Adverse Events (AE), Adverse Device Effects (ADE), Serious Adverse Events (SAE), and Serious Adverse Device Effects (SADE) were compared in participants receiving standard of care (SoC) alone versus the intervention group. The SoC group showed a slightly higher rate of AEs (1.43 events/100 patient days) compared to the intervention group (1.16 events/100 patient Days). ADEs were observed only in the intervention group at a rate of 1.00 events/100 patient days. No SAEs or SADEs were reported in either group.

### Surgical Wound Healing

By follow-up 1 (Day 14), 51% of participants in the intervention group had achieved wound healing, compared to 35% in the SoC group. These results are presented descriptively in Figure 7.

**Figure 7.**
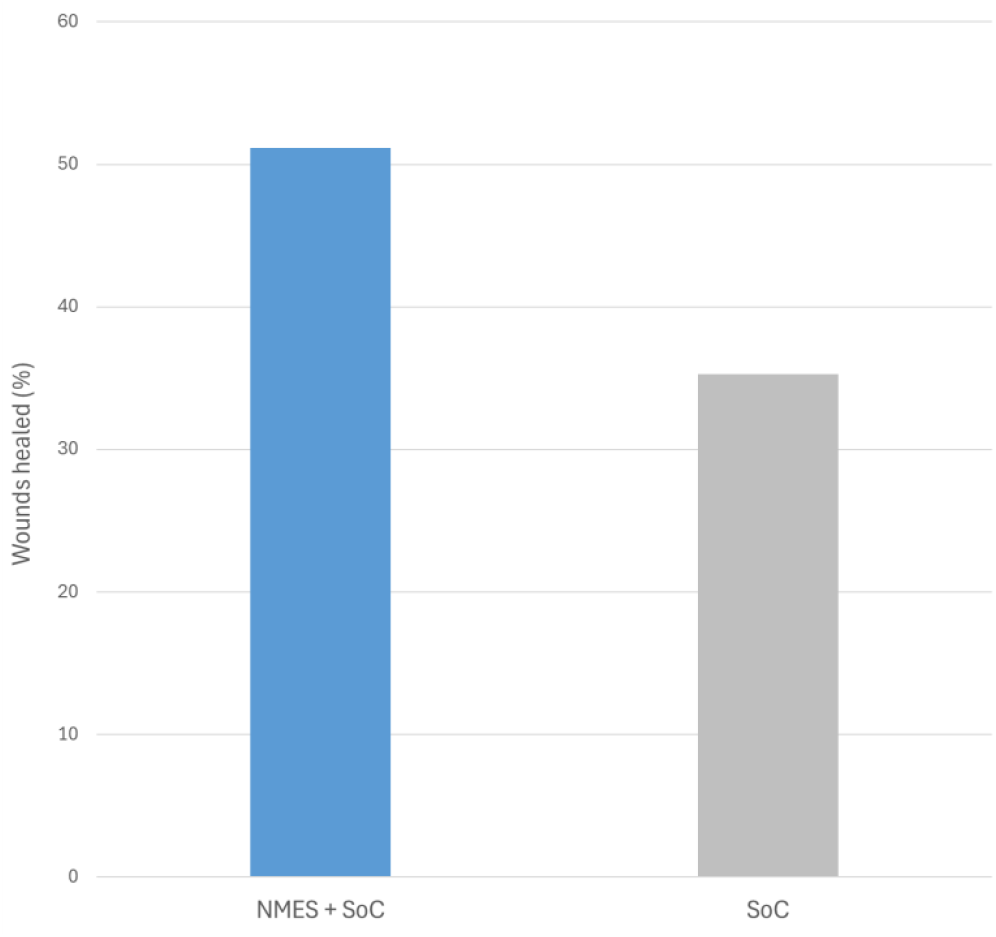
Proportion of Surgical Wounds Healed by Follow-up 1 (Day 14) in the Intervention and Standard Care (SoC) groups (N=78). 51% of participants in the intervention group achieved complete wound healing compared to 35% in the SoC group. The data are presented as the percentage of participants with fully healed wounds in each group.

### Pain

In the standard care group, pain scores measured by the visual analog scale (VAS) decreased from a postoperative mean (SD) of 41.2 (28.9) to a follow-up mean (SD) of 34.3 (24.3), representing a mean reduction of 6.9. In comparison, the intervention group showed a decrease from a postoperative mean (SD) of 39.0 (28.3) to a follow-up mean (SD) of 30.5 (22.1), representing a mean reduction of 8.5.

## Discussion

Exercise is frequently prescribed as an intervention for edema outside the orthopedic setting (28). Following foot and ankle surgery, however, patients are frequently advised not to bear weight on their operated limb (29). The rationale for this is to avoid mechanical damage to the healing structures. However, this goal conflicts with the goal of mitigating post-surgical edema, which is only one of several negative consequences of inactivity following surgery (30). Others include impairment to the cardiovascular system, and range of motion at the ankle (31). Additionally, the NMES device appeared to have a positive effect on subjects perceived pain and the rate healing at the surgical site.

Matching control and intervention groups proved challenging due to the inherent heterogeneity of the patient population. This was addressed by normalizing each subject’s involved limb using contralateral comparison and by evaluating edema trajectories relative to each subject’s immediate post-operative baseline.

Participants receiving NMES had 33% less edema (p<0.05) than those receiving standard care. Peak edema has passed by Day 14 (32), this suggests that the intervention group returns to a normative state more quickly than the SoC group and this reduction in edema could accelerate recovery time. The NMES device offers several of the advantages of a gentle exercise regimen for the lower limb, without the drawbacks of weight-bearing. The device stimulates the leg muscle pumps in a manner analogous to walking but activates multiple muscle groups simultaneously to achieve this without causing excessive movement. This action augments fluid flow out of the lower limb, and this study demonstrates that this effect is accompanied by significant reduction in swelling compared to SoC alone. This study further suggests that the device is safe and well tolerated in this patient population.

## Data Availability

All data produced in the present study are available upon reasonable request to the authors

## Acknowledgement

We gratefully acknowledge the involvement of Maidstone and Tunbridge Wells NHS Trust in supporting the conduct of the study.

